# Obtaining prevalence estimates of COVID-19: A model to inform decision-making

**DOI:** 10.1101/2020.08.06.20169656

**Authors:** Ida Sahlu, Alexander Whittaker

## Abstract

**Objectives:** The primary aim was to evaluate whether randomly sampling and testing a set number of individuals for active or past COVID-19 while adjusting for misclassification error captures a simulated prevalence. The secondary aim was to quantify the impact of misclassification error bias on publicly reported case data in Maryland.

**Methods:** Using a stratified random sampling approach, 50,000 individuals were selected from a simulated Maryland population to estimate the prevalence of active and past COVID-19. Data from the 2014-2018 and 2018 American Community Surveys were used. The simulated prevalence was 0.5% and 1.0% for active and past COVID-19, respectively. Bayesian models, informed by published validity estimates, were used to account for misclassification error when estimating the prevalence of active and past COVID-19.

**Results:** Failure to account for misclassification error overestimated the simulated prevalence for active and past COVID-19. Adjustment for misclassification error decreased the point estimate for active and past COVID-19 prevalence by 55% and 29%, respectively. Adjustment for sampling method and misclassification error only captured the simulated past COVID-19 prevalence. The simulated active COVID-19 prevalence was only captured when set to 0.7% and above. Adjustment for misclassification error for publicly reported Maryland data increased the estimated average daily cases by 8%.

**Conclusions:** Random sampling and testing of COVID-19 is needed but must be accompanied by adjustment for misclassification error to avoid over- or underestimating the prevalence. This approach bolsters disease control efforts. Implementing random testing for active COVID-19 may be best in a smaller geographic area with highly prevalent cases.

## Background

The population level prevalenceof COVID-19 at a specific point in time is unknown in the United States due to two important epidemiologic biases: selection bias and misclassification error bias. Selection bias arises because testing is generally restricted to those with the most common symptoms of or with known exposure to COVID-19.^1,2(p19)^ Misclassification error arises because the tests used are not perfect and may lead to false positives and false negatives.^3-6^ As a result, decisions on the re-opening of communities are being made without valid estimates on the extent to which COVID-19 is impacting communities at a specific time.^7^

Since not all COVID-19 cases are symptomatic, testing individuals who present with symptoms or who have known exposure to COVID-19 likely underestimates the true prevalence of COVID-19 in communities.^8,9^ Indeed, a study conducted in Iceland reported that only 57% of those confirmed with COVID-19 through random testing in the population had COVID-19 symptoms.^8^ Therefore, to address selection bias when obtaining population-level prevalence estimates of COVID-19, the reported testing results should derive from a randomly selected population as it helps identify potential asymptomatic individuals. It is especially important to identify asymptomatic individuals in the population because pre-symptomatic or asymptomatic individuals may transmit disease ^8,10,11^.

Additionally, current estimates of COVID-19 cases in the United States do not account for misclassification error that may arise due to the imperfect nature of the tests. Studies have reported that testing for active and past COVID-19 infection may lead to high false positives and negatives depending on the test and its associated validity estimates, defined by the sensitivity and specificity.^3,5^ Based on two meta-analyses, the sensitivity for the active COVID-19 infection diagnostic test with reverse transcriptase-polymerase chain reaction (RT-PCR) may be as high as 89% and as low as 63%.^3,4^ The specificity for this same test may be as high as 98.8%.^4^ Based on a meta-analysis of serology tests for past COVID-19 infection, the results varied depending on whether enzyme-linked immunoassay (ELISA) or Chemiluminescence Enzyme Immunoassays (CLIA) based methods were used. ELISA yields higher sensitivity and specificity values closer to 93.5% and 96.1%, respectively, compared to CLIA.^6^ With such varying validity estimates for active and past COVID-19 infection, misclassification error may be an important bias affecting current prevalence estimates.

Frequentist and Bayesian statistical approaches are available to account for test sensitivity and specificity when estimating the prevalence.^12,13^ One study conducted in Santa Clara, California found the prevalence of COVID-19 antibodies decreased by 20% from 1.5% (95%CI: 1.1-2.0%) to 1.2% (95%CI: 0.7-1.8%) after adjusting for the serological test validity estimates.^5^ Their adjustment applied a frequentist approach as opposed to a Bayesian approach, which has a couple disadvantages. The first disadvantage, as noted by the authors, is their model only holds if one minus the specificity is higher than the sample prevalence. This restriction is not needed for a Bayesian model. The second disadvantage is the frequentist approach uses point estimates for the test validity estimates, while the Bayesian approach allows for the validity estimates to vary by assigning a distribution. To our knowledge, Bayesian methods have yet to be applied in the published literature in the context of estimating active and past COVID-19 infection.

Ideally the number of new cases would be sufficient for decision-making, but this parameter is difficult to correctly estimate with asymptomatic cases. An alternative is to regularly obtain repeated population-level based prevalence estimates at specific points in time that account for selection and misclassification error biases. The primary aim of this study is to evaluate with a simulated population whether randomly sampling and testing a set number of individuals (i.e. the weekly average number of tests) for active or past COVID-19 while adjusting for misclassification error captures the simulated prevalence of active and past COVID-19 infection. As a secondary aim, we quantify the impact of misclassification error on publicly reported case report data. We use the state of Maryland as a worked example, but the methods implemented can be applied to any setting.

## Methods

### Simulation of Maryland Population

To create the simulated Maryland population from which to draw the study sample, we simulated the household, age and race distributions for each county in Maryland using the 2018 American Community Survey data for counties with more than 20,000 residents and the 2014-2018 American Community Survey data for counties with fewer than 20,000 residents. One-year data was not reported for smaller counties.

### Definition of outcomes

We define ‘active COVID-19’ as a positive result from any diagnostic test for the novel coronavirus, severe acute respiratory syndrome coronavirus 2 (SARS-CoV-2), collected from an individual’s respiratory system. Testing positive for SARS-CoV-2 either through reverse-transcription polymerase chain reaction (RT-PCR) or a point-of-care rapid diagnostic test (POC) would be considered active COVID-19. We define ‘past COVID-19 infection’ as an individual who would test positive for IgG, IgM and IgG, or total antibody through a serological test for a COVID-19 antibody response due to past exposure to SARS-CoV-2. We do not assign symptom status to individuals with active or past COVID-19 infection, given sampling was not dependent on the presence of symptoms.

### Assigning prevalence of active and past COVID-19 to simulated Maryland Populations

In the simulated populations, we assigned an average prevalence of 0.5% for active COVID-19 and 1% for past COVID-19, where the levels varied by county. These prevalence values are referred to as the simulated prevalence. To vary the active and past COVID-19 prevalence at the county level, we first grouped Maryland’s 24 counties into four groups based on the distribution of the cumulative number of cases reported by the Maryland Department of Health by June 10.^14^ These county groupings also aligned with the population distribution in the state, where more densely populated counties had a higher cumulative number of COVID-19 cases.^14^ We then assigned a prevalence value for each group as follows in descending order based on the number of reported COVID-19 cases: In Montgomery and Prince George’s counties, we assigned the prevalence of active and past COVID-19 at 1.0% and 2.0%, respectively. In Baltimore City and Baltimore County, we assigned the prevalence of active and past COVID-19 at 0.45% and 0.9%, respectively. In Anne Arundel, Carroll, Charles, Frederick, Harford, Howard, St. Mary’s, and Wicomico Counties, we assigned the prevalence of active and past COVID-19 at 0.25% and 0.5%, respectively. Finally, we assigned the prevalence of active and past COVID-19 at 0.06% and 0.1%, respectively, for the remaining counties. We believed these prevalence values were reasonable but likely low since the Maryland population is estimated at approximately 6,000,000 individuals and a minimum of approximately 50,000 individuals were diagnosed with COVID-19 by June 10,2020.^14,15^ We extended our analysis for active COVID-19 infection, however, to examine the change in results when the average prevalence was incrementally increased to 1% in the event our simulated prevalence is unrealistically low. If our methods accurately estimate small prevalence values, then then this approach can easily be applied to higher prevalence values. We note, however, it is truly impossible to determine the most realistic simulated prevalence since testing data reported to date primarily reflects those with more severe COVID-19 symptoms.

### Sampling of individuals for COVID-19 testing

To obtain a representative sample for Maryland, we randomly sampled 50,000 individuals (approximately 1%) separately for active and past COVID-19 from the simulated population using a stratified probability proportional-to-size approach. This sample size corresponds to approximately the average weekly number of tests reported in Maryland in May 2020.^14^ We used the counties of Maryland to partition the sampling frame into mutually exclusive exhaustive strata. This approach ensured individuals were selected from each county. To sample the 50,000 individuals, we first selected 50,000 households from the simulated population with representation from each stratum. The households’ selection probability was proportional to the household population size in the county. We then randomly selected a person from that household.

### Model Development

Four models were implemented to estimate the prevalence of active and past COVID-19 Model 1 reports the crude prevalence of active or past COVID-19 infection and does not account for selection or misclassification error biases. Model 2 accounts for the sampling design, which corrects for selection bias. Model 3 only accounts for misclassification error bias using a Bayesian approach. Model 4 combines Models 2 and 3 to account for both types of biases. We describe in detail each model below.

Model 1 was a crude estimate of the proportion of those with active or past COVID-19, which divided the number of positive tests by the total sample size. To compare Model 1 to the models using a Bayesian approach, we calculated a Bayesian credibility interval (BCI) for Model 1 using an uninformative Beta(l,l) prior on the prevalence.

Model 2 accounted for the sampling design and reduced selection bias. For each individual, we obtained their selection probability by multiplying the probability of their household being selected in the county by their probability of being selected from within the household. The inverse of the individual’s selection probability was used as the weight. Specifically, Model 2 estimated the prevalence of both outcomes separately as follows,

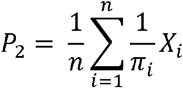

where, *n* is the sample size, *π* is the individual’s selection probability and *X_i_* is a binary variable being 1 if the individual tested positive and 0 if they tested negative. The variance of this estimate was calculated with replicate survey weights with the finite population correction applied to account for the sample being a small fraction of the total population. We reported 95%Clfor binomial proportions for this model.

Model 3 accounted for misclassification error (false positives and false negatives) due to the test accuracy. We used a Bayesian model which allows to encode information using prior distributions on test accuracy into the model and allows test accuracy to vary.^12^. Mean and 95%CI in published literature were used to inform the prior distributions on test sensitivity and specificity for both active and past COVID-19.^3-6^ Specifically, for active COVID-19, we used the pooled sensitivity value by Kim *et al*. and specificity value by Zitek.^3,4^ The following prior distributions of

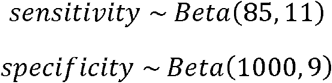

were used corresponding to a mean and 95% Cl for sensitivity at 89.0% (95%CI: 81.0% - 94.0%) and for specificity at 99.0% (95%CI: 98.5 - 99.5%). Specifically for past COVID-19, we used the Kontou *et al*. sensitivity estimates and the Bendavid, E *et al*. specificity estimates.^5,6^ The following prior distributions of

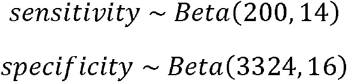

were used corresponding to a mean and 95%CI for sensitivity at 93.5% (95%CI: 90.0 - 97.0%) and for specificity at 99.5% (95%CI: 99.2% - 99.7%)^5,6^. Model 3 estimated prevalence as follows:

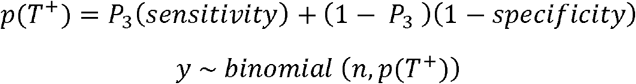

where *p*(*T*^+^) is the probability of testing positive, *P*_3_ is the population prevalence, *n* is the sample size, and *y* is the number of individuals with active or past COVID-19. We reported the posterior means and the 95% BCI of *P*_3_ for both outcomes. We calculated the posterior distribution using the No-U-Turn Sampler, a variant of Hamiltonian Monte Carlo.^16^ We ran four Markov Chains with 4,000 iterations of warmup and 9,000 iterations total. The No-U-Turn sampler has one hyperparameter, maximum tree depth, which was set to 30. Four chains are the default for the algorithm. The tree depth was adjusted until the algorithm converged.

Model 4 accounted for the sampling design and misclassification error by combining Models 2 and 3. Model 4 used jackknife splitting to account for the survey weight and misclassification error bias. As described by Kang and Bernstein and Cao *et al*. on how to combine survey methodology with Bayesian modeling^17,18^, we split the sample data using leave one stratum out jackknife splits and estimated Model 3 on the jackknife split. This was done for each stratum, generating 24 prevalence estimates (one for each county), which were weighted by the size of the stratum left out of the split and averaged with the stratum weights. Specifically,

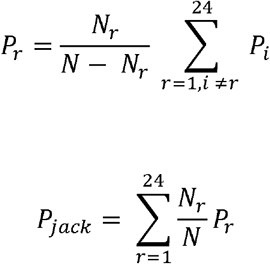

where *P_i_* is the Model 3 estimate calculated with the *i*th stratum left out, *P_r_* is the weighted stratum estimate, *N_r_* is the left out stratum size, *N* is the population size and *P_jack_* is the jackknife estimate of the prevalence. The variance of the jackknife estimates variance is

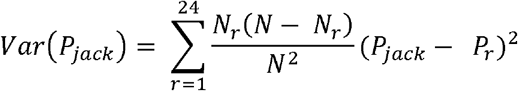

We reported *P_jack_* and 95% CIs for binomial proportions calculated using *Var(P_jack_)*.

### Evaluating the impact of correcting for sampling and misclassification error

To examine misclassification error independently of sampling error, we compared Model 1 to Model 3, and Model 2 to Model 4. To appropriately compare Models 1 and 3, we obtained point estimates and 95% BCI. We calculated point estimates and 95% CIs for Models 2 and 4 since these are frequentist estimation techniques.

To quantify the impact of misclassification error on publicly reported case report data we first estimated the crude percent positive tests for each day between May 6^th^ and May 26^th^, 2020 reported by the Maryland Department of Health’s COVID-19 dashboard.^14^ To do so, we divided the number of new cases reported by the total number of tests conducted. The total number of tests conducted was obtained by summing the number of new cases and the number of negatives tests on that date. Thereafter, we ran Model 3 with the prior distributions for active COVID-19 infection test validity estimates to account for potential false positives and false negatives.

All data were analyzed using RStudio Version 1.2.5042 and R Foundation for Statistical Computing 4.0.0. The rstan package, the interface to Stan in R, was used for the Bayesian models. The R survey package was used to calculate the estimates for Model 2.^19^

## Results

### Study sample

A total of 50,000 (0.9%) simulated individuals were sampled from the 2,213,911 households in the simulated Maryland population. Table 1 describes the population characteristics of the study sample. Females represented 51.0% of the population, over 50% were between the ages of 20 and 49 years, and the four counties with the highest representation were Montgomery (16.6%), Prince George’s (14.2%), Baltimore City (10.7%) and Baltimore County (14.2%).

**Table 1:** Distribution of population characteristics among the simulated Maryland study sample (n=50,000)

| Sample population characteristics (N=50,000) | N(%) |
| --- | --- |
| **Sex** |  |
| Female | 25525 (51.0%) |
| Male | 24475 (49.0%) |
| **Race/Ethnicity** |  |
| White alone | 26224 (52.4%) |
| Black or African American alone | 14164 (28.3%) |
| Hispanic or Latino | 4791 (9.6%) |
| Asian alone | 2926 (5.9%) |
| Other | 293 (3.8%) |
| **Age groups** |  |
| 0-9 | 1445 (2.9%) |
| 10-19 | 4017 (8.0%) |
| 20-29 | 7576 (15.2%) |
| 30-39 | 10723 (21.4%) |
| 40-49 | 10963 (21.9%) |
| 50-59 | 7988 (16.0%) |
| 60-69 | 4350 (8.7%) |
| 70-79 | 1791 (3.6%) |
| 80+ | 1147 (2.3%) |
| **County residence** |  |
| Allegany County | 616 (1.2%) |
| Anne Arundel County | 4809 (9.6%) |
| Baltimore city | 5356 (10.7%) |
| Baltimore County | 7079 (14.2%) |
| Calvert County | 729 (1.5%) |
| Caroline County | 271 (0.5%) |
| Carroll County | 1367 (2.7%) |
| Cecil County | 829 (1.7%) |
| Charles County | 1293 (2.6%) |
| Dorchester County | 304 (0.6%) |
| Frederick County | 2172 (4.3%) |
| Garrett County | 275 (0.6%) |
| Harford County | 2141 (4.3%) |
| Howard County | 2632 (5.3%) |
| Kent County | 176 (0.4%) |
| Montgomery County | 8307 (16.6%) |
| Prince George's County | 7118 (14.2%) |
| Queen Anne County | 408 (0.8%) |
| Somerset County | 189 (0.4%) |
| St. Mary's County | 932 (1.9%) |
| Talbot County | 379 (0.8%) |
| Washington County | 1270 (2.5%) |
| Wicomico County | 854 (1.7%) |
| Worcester County | 494 (1.0%) |
| **Number of individuals in household** | 2.58 (1.00, 10.0) |

Table 2 describes the prevalence estimates obtained from the four models for both active and past COVID-19 infection by sample size. When we sampled 50,000 individuals, the crude prevalence estimates obtained through Model 1 for both active and past COVID-19 infection did not capture the simulated Maryland prevalence. Once we adjusted for misclassification error in Model 3, the point estimates for the prevalence of active and past COVID-19 infection decreased by 55% and 29%, respectively, from Model 1. When we only accounted for the sampling approach only (Model 2), we did not capture the simulated prevalence for active and past COVID-19 infection. When we adjusted for misclassification error in Model 4, the point estimate for active and past COVID-19 prevalence decreased by 60% and 41%, respectively, from Model 2 estimates. Therefore, the prevalence estimates for active and past COVID-19 infection decreased when we accounted for misclassification error. When comparing the prevalence estimates to the simulated prevalence, only Model 3 captured the simulated prevalence for both active and past COVID-19. Model 4 only captured the simulated prevalence for past COVID-19 infection.

**Table 2:** Prevalence estimates for active and past COVID-19 infection for each model type for the50,000 individuals sampled from the simulated Maryland population.

|  | N=50,000 |  |
| --- | --- | --- |
| Model type | Prevalence estimate for active COVID-19, where simulated prevalence is 0.5% | Prevalence estimate for past COVID-19, where simulated prevalence is 1.0% |
| Model 1: Crude (No adjustment) | 1.4% (95% BCI: 1.3%, 1.5%) | 1.4% (95% BCI: 1.4%, 1.5%) |
| Model 2: Adjustment for sampling error without adjustment for misclassification error | 1.5% (95% CI: 1.4%, 2.0%) | 1.7% (95% CI: 1.6%, 2.0%) |
| Model 3: Adjustment for misclassification error without adjustment for sampling error | 0.6% (95% BCI: 0.1%, 1.2%) | 1.0% (95% BCI: 0.7%, 1.3%) |
| Model 4: Combines adjustment for sampling and misclassification error | 0.6% (95% CI: 0.6%, 0.7%) | 1.0% (0.8%, 1.1%) |

Figure 1 illustrates how the estimated prevalence of active COVID-19 changes when the average simulated population prevalence increases from 0.5% to 1%. When the simulated active COVID-19 prevalence is at 0.7% or above, then Model 4 is able to capture the simulated population’s prevalence.

**Figure 1:**
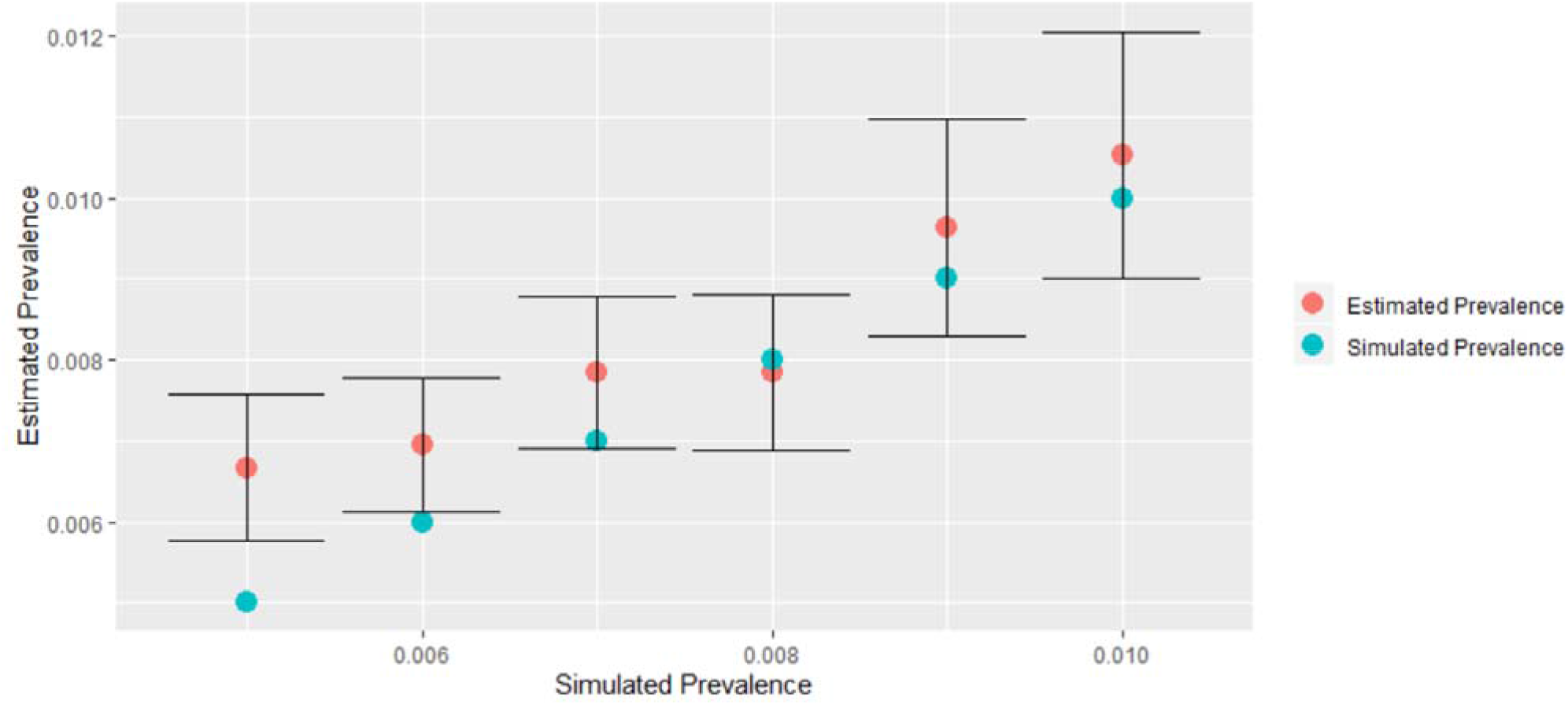
Estimated active COVID-19 Infection prevalence when testing 50,000 individuals using Model 4, which accounts for sampling design and misclassification error, for simulated prevalence values ranging from 0.5% to 1%.

### Analysis of reported Maryland COVID-19 cases

Between May 6^th^ and May 26^th^, the average daily number of new cases reported by the Maryland Health Department was 979.5. Once we accounted misclassification error, the estimated average daily number of new cases increased by 8% for that same time period. Figure 2 illustrates the daily percent of positive active COVID-19 cases before and after adjustment for misclassification error. Failing to account for misclassification error (false positives and false negatives) generally leads to an increase in the active COVID-19 cases. Figure 3 demonstrates the cumulative effect of failing to account for diagnostic error for the active COVID-19 diagnotic test, which is the difference between the adjusted and unadjusted daily percent positive cases. We estimate by May 26^th^ approximately 1811 individuals may have been misclassifed as not having active COVID-19.

**Figure 2:**
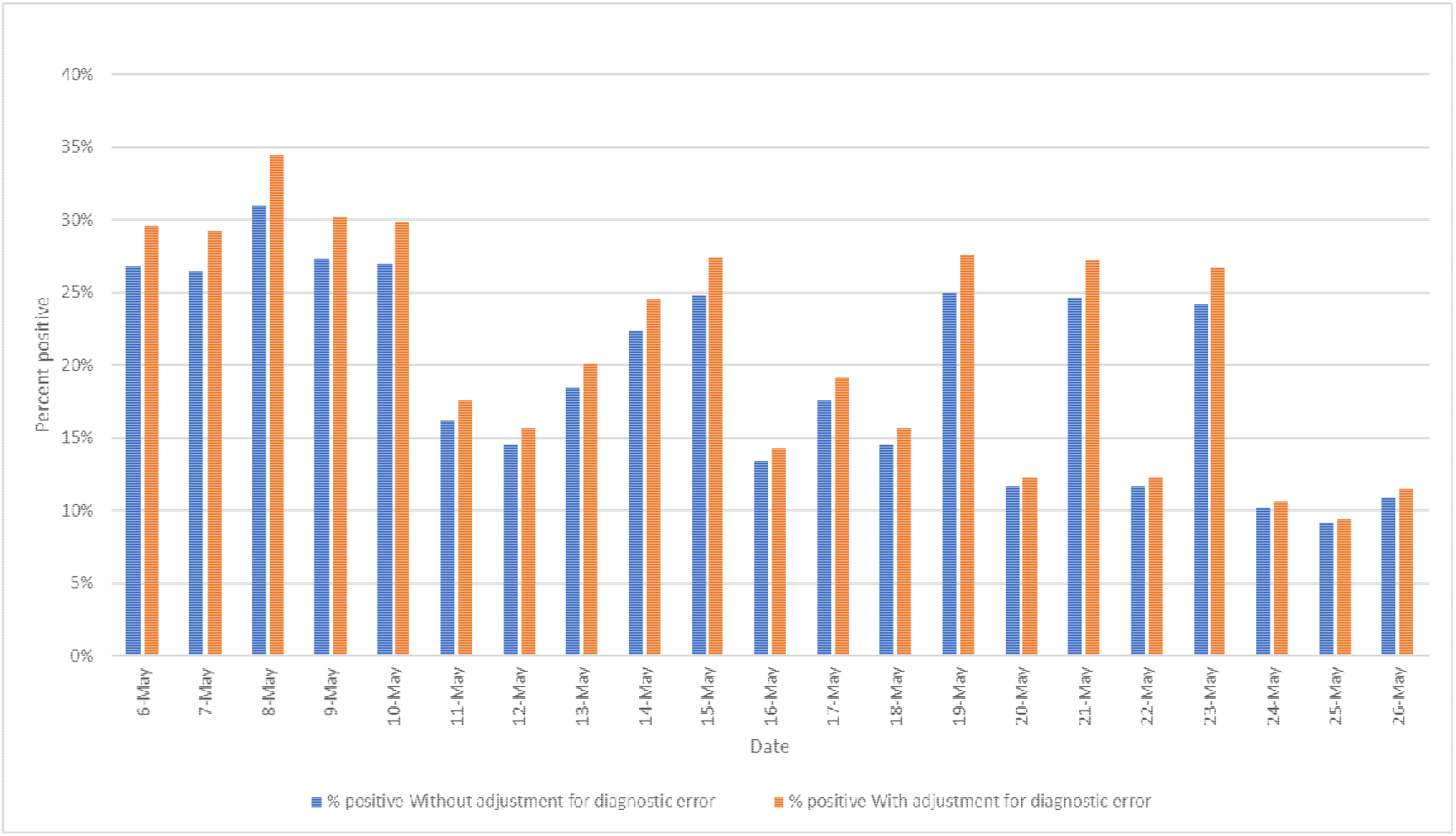
Percent positive active COVID-19 infection cases with and without adjustment for diagnostic error for Maryland between May 6th and May 26th, 2020.

**Figure 3:**
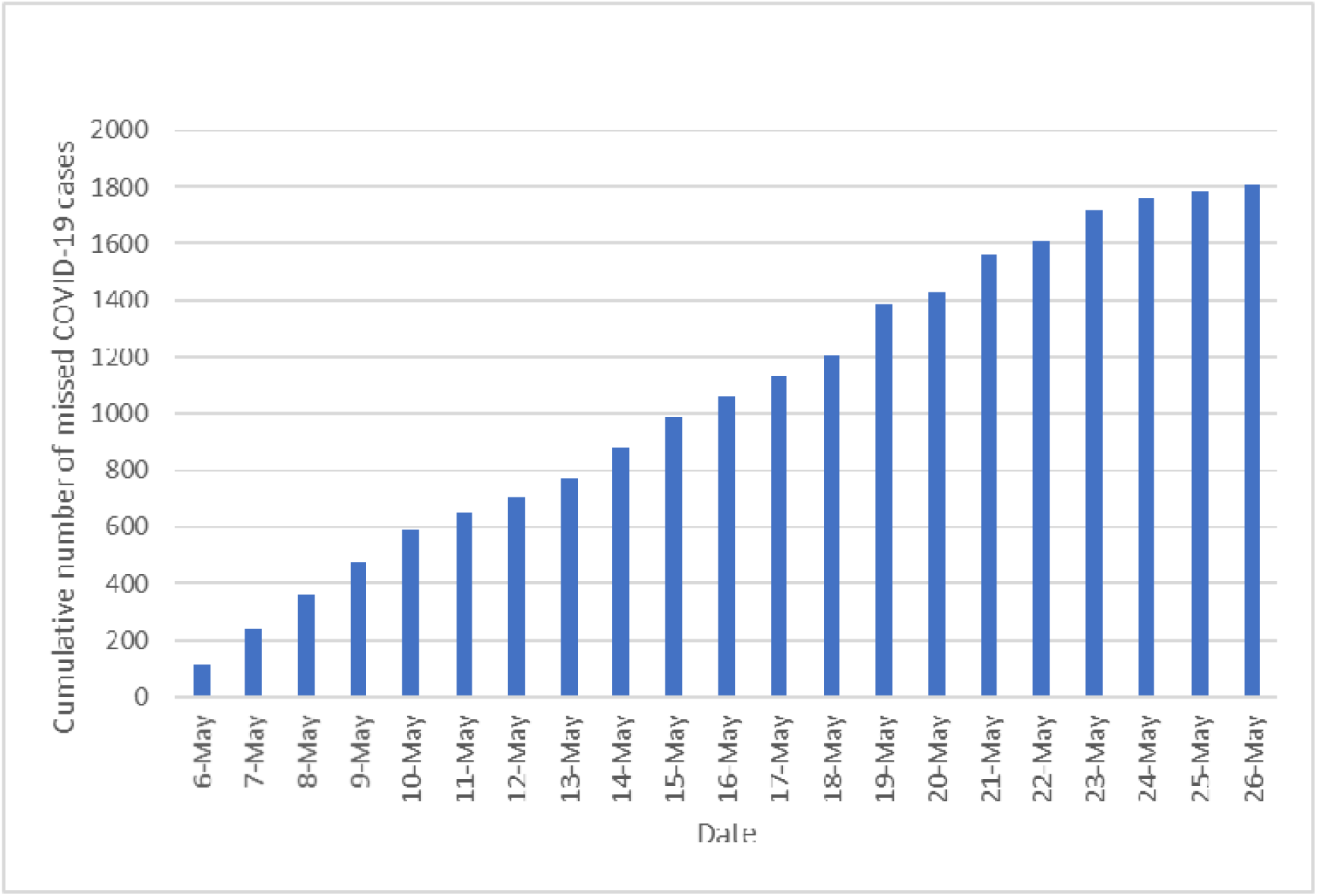
Estimated cumulative number of missed cases in Maryland between May 6th and May 26^th^, 2020.

## Discussion

Overall, randomly sampling and testing individuals from the population combined with adjustment for misclassification error captured the simulated prevalence of active and past COVID-19 for a set number of tests (i.e. the weekly average) depending on the population’s underlying prevalence. Adjustment for misclassification error alone led to a decrease in the estimated population-level prevalence by 55% and 29% for active and past COVID-19, respectively, thereby showing the importance of false positives and false negatives. When we simultaneously adjusted for the sampling scheme and misclassification error, the estimated prevalence confidence intervals only captured the simulated past COVID-19 prevalence. We captured the simulated active COVID-19 prevalence once the simulated prevalence was increased from 0.5% to 0.7% given the diagnostic test accuracy. Failure to capture the simulated active COVID-19 prevalence at 0.5% may be explained by the limited variation in the county-specific prevalence values, hence the narrow 95%CI. These findings are unsurprising given prevalence estimates are more sensitive to diagnostic error and prone to having false positive results when the prevalence is low.^19,20^ Our methodology improved on Bendavid, E. *et al*’s seroprevalence study which accounted for misclassification error^5^ in that we randomly selected individuals for testing from the simulated population. As a result, our estimates were not subject to selection bias and were a direct representative sample of the population. Bendavid, E. *et al* did attempt to account for selection bias by using post-stratification weights. However, adjustment is only as good as the variables included in the post-stratification weights and, therefore, it is difficult to assess whether their adjustment was successful. Finally, we employed a Bayesian approach to allow testing sensitivity and specificity to vary, which was a more realistic assumption compared to the frequentist approach used by Bendavid, E. *et al*. Since various tests are available for diagnosis, it is especially important to allow the sensitivity and specificity values used as priors in the model to vary.

We also found misclassification error to be an important bias for publicly reported cases. When we examined the impact of misclassification error on publicly reported COVID-19 cases in Maryland between May 6^th^ and May 26^th^, 2020, adjustment for false positives and false negatives led to higher prevalence estimates than previously reported. The increase in the prevalence estimate, albeit small, meant the number of false negatives exceeded the number of false positives. We demonstrated the cumulative effect of daily missed active COVID-19 cases may lead to approximately 1811 individuals being misclassified in the 20 days studied. This means failure to appropriately diagnose and isolate these 1811 individuals could lead to an additional 1,594 (=1811*0.88) secondary cases in Maryland if the reproductive number of active COVID-19 infection at the end of the time period (May 26^th^, 2020) was 0.88 as reported by rt.live^21^. The importance of false negatives in our findings is unsurprising since false negatives are of greater concern than false positives when the prevalence is high.^20,22^ In this case, the daily percent positive reported by the Maryland Department of Health during that time period was greater than 10% and could be considered high. If the percent positive values were lower, for example closer to the simulation prevalence values of 1%, then false positives would be more of a concern. Therefore, although the direction of bias due to misclassification error was different from the simulated data, the results were not surprising. Our findings allowed us to confirm misclassification error presents a problem for already collected data and adjustment is needed to minimize bias.

There are some limitations in our study. First, our study used simulated data to randomly select individuals and estimate the prevalence of active and past COVID-19 in Maryland while adjusting for misclassification error. As a result, our analysis does not reflect some additional biases that may arise in the field, such as non-response bias from individuals refusing to be tested. Any implementation of our proposed sampling and analysis approach in the field would need further adjustment for non-response bias. The advantage, however, of simulated data was our ability to determine whether the random sampling of individuals followed by adjustment for misclassification error bias in the absence of other biases captured our simulated prevalence of active and past COVID-19. This level of comparison would not be feasible with real data. Second, our simulated prevalence of 0.5% and 1% for active and past COVID-19, respectively, may not be correct for Maryland. Since it is impossible to validate these values given the true prevalence is unknown, we decided to choose low, yet, realistic prevalence values to implement this study. Third, we did not know which exact tests are most generally used in Maryland to diagnose individuals so we used published sensitivity and specificity values to adjust our model. Therefore, our results should be interpreted with caution and the data should be re-analyzed with the sensitivity and specificity values specific to the diagnostic tests used if available. We did, however, allow our sensitivity and specificity values to vary with the Bayesian approach and likely captured the validity estimates of the tests used in Maryland. Finally, we could not guarantee the information reported by the Maryland Department of Health did not count individuals more than once. For example, some healthcare providers might be attempting to account for misclassification error directly in the field by conducting two subsequent tests on one individual. This was a possibility because the U.S. Centers for Disease Control and Prevention states in the context of antibody testing that an orthogonal testing algorithm may be employed where two independent tests are administrated to minimize false positives.^23^ Since we examined active COVID-19 as opposed to past COVID-19 and our results were unsurprising, we do not believe this is an important concern for our estimates. It should also be noted the Maryland Department of Health reports new cases as opposed to new test results.

In summary, through the random sampling and testing of individuals in our simulated Maryland population, we recovered the simulated prevalence of active and past COVID-19 after accounting for misclassification error. Our analysis demonstrated random sampling of individuals from the general population is needed to truly understand the extent to which populations are impacted by COVID-19 at a specific point in time. However, random sampling must be accompanied by adjustment for misclassification error to obtain valid estimates, otherwise failure to account for this bias may lead to an over- or underestimate of active or past COVID-19. The added benefit of implementing random testing in the community is that it helps with the identification and isolation of asymptomatic cases and bolsters contact tracing efforts. We found capturing the prevalence of active COVID-19 may be difficult on a large scale if the prevalence is too low. Therefore, it may be best to implement such a study in a smaller geographic area with more prevalent cases, such as at the county level. If large-scale testing of individuals is not possible due to budget constraints, testing costs could be reduced by testing pooled samples.^24^ This approach could reduce costs since fewer tests are implemented when trying to obtain population-level prevalence estimates. We recommend future research examine the feasibility of implementing such an approach. Finally, although we conducted our analysis using a simulated population of Maryland, this methodology can be employed in any state or municipality. The statistical model can be adapted to the setting by modifying the priors on test sensitivity and specificity, and if available, on population prevalence. We believe government and community leaders should implement the approach described in this study to ensure they make decisions about the re-opening and potential future re-closing of communities based on valid data. Without such information, decisions will be made with biased data and without the knowledge of whether the presence of COVID-19 is truly low enough to resume regular activities. If communities are re-opened too prematurely or closed too late, specifically when there are untested individuals asymptomatic COVID-19 cases, then there may be a re-emergence of the pandemic.

## Data Availability

Data are not publicly available but can be requested by contacting the corresponding author.

## Notes

### Competing Interest Statement

The authors have declared no competing interest.

### Funding Statement

This work was supported by the MITRE Corporation. The authors did not receive external funding for this work.

### Author Declarations

This work was approved by the MITRE Corporation's Information Release Office. This study does not constitute human subjects research, and therefore, IRB approval or exemption was not needed.

### Summary of Updates

Revised typo in Abstract and Results sections of manuscript.

